# COVID-19 mortality rate in Russia: forecasts and reality evaluation

**DOI:** 10.1101/2020.09.25.20201376

**Authors:** Lifshits Marina, Neklyudova Natalia

**Affiliations:** Institute of Economics, Ural Branch of the Russian Academy of Sciences, Russia

**Keywords:** COVID-19, regions of Russia, mortality rate, lethality, econometric modeling

## Abstract

**Relevance:** COVID-19 is an extremely dangerous disease that not only spreads quickly, but is also characterized by a high mortality rate. Therefore, predicting the number of deaths from the new coronavirus is an urgent task.

**Research objective:** The aim of the study is to analyze the factors affecting COVID-19 mortality rate in various countries, to predict direct and indirect victims of the pandemic in the Russian Federation, and to estimate additional mortality during the pandemic based on the demographic data.

**Data and methods:** The main research method is econometric modeling. Comparison of various data was also applied. The authors’ calculations were based on data from the RSSS, the World Bank, as well as specialized sites with coronavirus statistics in Russia and in the world.

**Results:** A predictive estimation of the deceased number of people due to the pandemic in Russia was made. It is confirmed that the deaths proportion of the completed cases of the disease depends on the level of testing. It is shown that the revealed mortality of the disease depends on the proportion of completed cases, on the population age structure, and on how early the pandemic entered the country compared to the other countries. It is determined that the number of additional deaths due to the coronavirus is approximately 31 thousand people.

**Conclusions:** The analysis revealed that the relatively low proportion of COVID in Russia is the result of a special approach to the cause of death determination. The mortality rate in Russia in April 2020 was about 3% higher than in April 2019. The share of the deceased health workers in the total coronavirus mortality in the Russian Federation is higher than in the developed countries, which indicates an underestimation of the data on COVID-19 deaths in the Russian Federation, and the unsatisfactory quality of the Russian healthcare system. The number of direct and indirect victims of the pandemic in the Russian Federation at the end of July was approximately 43 thousand people.

## Introduction

The COVID-19 outbreak began in Russia a month later than in Italy and Spain, but the negative scenario was not avoided, the experience of Germany and some other countries did not help. By the end of April, Russia became number 9 among the countries with the highest number of cases (overtaking China), starting from 17-20 May, the Russian Federation was rated second, surpassed later by Brazil, and then India. Starting from July 5th, Russia ranked number 4 (table 1). During May Russia ranks second behind the US for active cases (all cases minus diseased and recovered), and from mid-April to mid-May also was second after the US for new weekly cases.

**Table 1.** Leaders in the total number of people infected with new coronavirus at the end of July and their rank as of the specified date (end of the day)

| Country | April<br>27 | May<br>07 | May<br>17 | May<br>27 | June<br>07 | June<br>17 | June<br>27 | July<br>07 | July<br>17 | July<br>27 |
| --- | --- | --- | --- | --- | --- | --- | --- | --- | --- | --- |
| USA | 1 | 1 | 1 | 1 | 1 | 1 | 1 | 1 | 1 | 1 |
| Brazil | 11 | 8 | 5 | 2 | 2 | 2 | 2 | 2 | 2 | 2 |
| India | 17 | 14 | 11 | 10 | 6 | 4 | 4 | 3 | 3 | 3 |
| Russia | 9 | 5 | 2 | 3 | 3 | 3 | 3 | 4 | 4 | 4 |
| South Africa | 51 | 44 | 39 | 30 | 23 | 21 | 18 | 14 | 6 | 5 |
| Mexico | 25 | 20 | 17 | 17 | 14 | 14 | 11 | 9 | 8 | 6 |
| Peru | 16 | 13 | 12 | 12 | 8 | 7 | 7 | 5 | 5 | 7 |
| Chile | 28 | 24 | 19 | 15 | 13 | 9 | 8 | 6 | 7 | 8 |
| Spain | 2 | 2 | 3 | 4 | 4 | 6 | 6 | 7 | 9 | 9 |
| UK | 6 | 4 | 4 | 5 | 5 | 5 | 5 | 8 | 10 | 10 |
| Iran | 8 | 10 | 10 | 11 | 10 | 10 | 10 | 10 | 11 | 11 |
| Pakistan | 27 | 22 | 20 | 18 | 16 | 15 | 12 | 12 | 12 | 12 |
| Saudi Arabia | 22 | 17 | 16 | 16 | 15 | 16 | 15 | 13 | 13 | 13 |
| Colombia | 48 | 42 | 38 | 32 | 27 | 23 | 21 | 19 | 18 | 14 |
| Italy | 3 | 3 | 6 | 6 | 7 | 8 | 9 | 11 | 14 | 15 |
| Turkey | 7 | 9 | 9 | 9 | 11 | 12 | 13 | 15 | 15 | 16 |
| Bangladesh | 45 | 37 | 30 | 23 | 20 | 18 | 17 | 18 | 17 | 17 |
| Germany | 5 | 7 | 8 | 8 | 9 | 11 | 14 | 16 | 16 | 18 |
| France | 4 | 6 | 7 | 7 | 12 | 13 | 16 | 17 | 19 | 19 |
| Argentina | 53 | 54 | 50 | 44 | 38 | 34 | 27 | 23 | 20 | 20 |

Compiled by the authors based on the data from https://www.worldometers.info/coronavirus/

The list of leaders in the total number of COVID-19 -related deaths over the last two months have not changed as much as the total number of people infected, since the mortality in the first European countries affected with the new disease was particularly high, besides the growth in the number of infections over time is reflected in the increase in the number of deaths (table 2). Differences in approaches to the statistical view of the number of infected and number of deaths affected the difference in the ranks, more on that later.

**Table 2.** Leaders in the total number of deaths due to COVID-19 at the end of July and their rank as of the specified date (end of the day)

| Country | April<br>27 | May<br>07 | May<br>17 | May<br>27 | June<br>07 | June<br>17 | June<br>27 | July<br>07 | July<br>17 | July<br>27 |
| --- | --- | --- | --- | --- | --- | --- | --- | --- | --- | --- |
| USA | 1 | 1 | 1 | 1 | 1 | 1 | 1 | 1 | 1 | 1 |
| Brazil | 11 | 6 | 6 | 6 | 3 | 2 | 2 | 2 | 2 | 2 |
| UK | 5 | 2 | 2 | 2 | 2 | 3 | 3 | 3 | 3 | 3 |
| Mexico | 16 | 15 | 12 | 9 | 7 | 7 | 7 | 5 | 4 | 4 |
| Italy | 2 | 3 | 3 | 3 | 4 | 4 | 4 | 4 | 5 | 5 |
| India | 19 | 16 | 16 | 14 | 12 | 8 | 8 | 8 | 8 | 6 |
| France | 4 | 5 | 4 | 4 | 5 | 5 | 5 | 6 | 6 | 7 |
| Spain | 3 | 4 | 5 | 5 | 6 | 6 | 6 | 7 | 7 | 8 |
| Peru | 21 | 19 | 18 | 17 | 15 | 14 | 11 | 10 | 10 | 9 |
| Iran | 8 | 9 | 9 | 10 | 10 | 10 | 9 | 9 | 9 | 10 |
| Russia | 20 | 20 | 19 | 18 | 14 | 13 | 13 | 11 | 11 | 11 |
| Belgium | 6 | 7 | 7 | 7 | 8 | 9 | 10 | 12 | 12 | 12 |
| Germany | 7 | 8 | 8 | 8 | 9 | 11 | 12 | 13 | 13 | 13 |
| Chile | 41 | 37 | 36 | 28 | 20 | 20 | 16 | 15 | 15 | 14 |
| Canada | 13 | 12 | 10 | 11 | 11 | 12 | 14 | 14 | 14 | 15 |
| Colombia | 35 | 33 | 31 | 30 | 27 | 24 | 22 | 22 | 16 | 16 |
| South Africa | 54 | 50 | 45 | 35 | 31 | 27 | 25 | 23 | 22 | 17 |
| Netherlands | 10 | 10 | 11 | 12 | 13 | 15 | 15 | 16 | 17 | 18 |
| Pakistan | 32 | 28 | 26 | 24 | 21 | 21 | 21 | 20 | 19 | 19 |
| Sweden | 14 | 14 | 15 | 16 | 17 | 16 | 17 | 17 | 18 | 20 |

Compiled by the authors based on the data from https://www.worldometers.info/coronavirus/

In April, COVID-19 seemed to be an extremely dangerous disease, not only quick spreading (the weekly number of the new cases in the world is still growing and in the last week of July reached its maximum), but also with a high mortality rate. On April 10th-12th, the share of deaths among the registered cases in the world exceeded 22%, but gradually declined and by the end of July dropped below 6%, but even in recent months, the mortality rate still remains ten times higher than from influenza. Therefore, predicting the number of deaths from the new coronavirus was a very urgent task in April and May.

A number of modelling and forecasting tools quantifying the future COVID-19 burden are available in different countries (Harapan et al., 2020; Haghani et al., 2020). Sebastiani et al. (2020) described the trends of Covid-19 spared in Italy. Anastassopoulou et al. (2020) and Gao et al. (2020) revealed the potential total numbers of COVID-19deaths in China. Shojaee et al. (2020) estimated the rate of death in Italy, Iran and South Korea. Semenova et al. (2020) made prognoses on due to CoVID deaths in Kazakhstan. Gerli et al. (2020) tried to forecast mortality trends in the 27 countries of the European Union, Switzerland and the United Kingdom. Al-Raeei (2020) calculated the coefficient of mortality of the COVID-19 pandemic for China, the United States, Russia, and the Syrian Arab Republic. Zemtsov & Baburin (2020) described situation in Russia.

The first COVID-19 mortality projections in Russia were calculated by the authors according to the data from the other countries as of May 7^th^ (Lifshits, 2020). Later, the authors gave an estimation of the “additional” mortality during the pandemic based on the FSSS (Federal State Statistic Service) demographic data for April^2^. This work also presents the authors’ calculations based on the FSSS data for May.

### Methodology and data

The main research method is econometric modeling. Comparison of various data was also applied. In addition to the FSSS data^3^, the paper used information from the World Bank (for the population age structure in the other countries)^4^, as well as specialized Russian^5^ and world websites^6^ with the coronavirus statistics.

## Results and Discussion

### COVID-19 mortality forecast in the Russian Federation based on the data from the countries of the world

The econometric models for COVID-19 mortality predictions in Russia were based on data from 107 countries and territories out of 208, which were selected according to the following criteria: the total number of cases at least 450 (by the end of May 7th, there were 122 such countries); the Worldometer website has data on testing (9 dropped out) and the number of recoveries (3 more removed); the World Bank website contains data on the population age structure (3 more are excluded from consideration).

The most significant disadvantage of the used statistics is that the COVID-19 mortality data from different countries are not comparable (Danilova, 2020). In some, as in Italy, all dead infected people are classified as COVID-19 victims, in others - criteria are different. Although the scientific research shows that the new coronavirus causes non respiratory disease, its mechanism is much more complex (Varga et al., 2020), therefore, Italy’s approach is more correct. On April 16th WHO just published recommendations, according to which “a death due to COVID-19 is defined as a death resulting from a clinically compatible illness, in a probable or confirmed COVID-19 case, unless there is a clear alternative cause of death (e.g. trauma). There should be no period of complete recovery from COVID-19 between illness and death. A death due to COVID-19 may not be attributed to another disease (e.g. cancer) and should be counted independently of preexisting conditions that are suspected of triggering a severe course of COVID-19”^7^. However, the interpretation of these words is different in different countries. Moreover, it should be noted that before the publication of these recommendations, 148 thousand deaths from COVID-19 out of 270 thousand were already registered in the world. The econometric models described below were built on the basis of that data, though each country followed its own rules.

The approach to the statistical calculation of the recoveries in different countries also varies. The UK, the Netherlands and Norway do not publish these numbers at all, so we had to exclude these countries from our econometric study. In some other countries, on the other hand, there may be too much rush to declare sick people recovered.

However, even with these important limitations, the econometric modeling can trace some patterns in COVID-19 mortality rate (*proportion of deaths from completed cases*) in different countries. That indicator in percentage is taken as an explainable variable in models 1 and 2 (Table 3).

**Table 3.** Regressors explaining the differences in COVID-19 mortality in different countries

| Regressors | Model 1 |  | Model 2 |  |
| --- | --- | --- | --- | --- |
|  | Coefficients | Std. Error | Coefficients | Std. Error |
| Constant | 21,36**** | (3,76) | 22,19**** | (3,76) |
| Tests/Infected | -0,05070** | (0,02398) | -0,04386* | (0,02412) |
| Completed cases, % | -0,1995**** | (0,0347) | -0,1934**** | (0,0345) |

<sup>7</sup> https://www.who.int/classifications/icd/Guidelines_Cause_of_Death_COVID-19.pdf

|  |  |  |  |  |
| --- | --- | --- | --- | --- |
| 65+/15+ | 0,3233*** | (0,1224) | 0,3452*** | (0,1220) |
| Data_250 | -0,1403** | (0,0649) | -0,1581** | (0,0651) |
| Tests/Population |  |  | -0,05395* | (0,03209) |
| Adj. R2 | 0,351 |  | 0,362 |  |
| N | 107 |  | 107 |  |

Table 3 shows the coefficients *b*_*i*_ of econometric equations

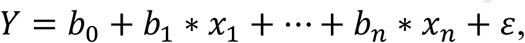

where *Y* is the explained variable, *x*_*i*_ is regression, *ε* is the remainder of the equation.

The following designations are stated in the table: Constant - free term of the equation; Tests / infection - the number of tests per diagnosed infected person; Completed, % - the percentage of completed cases (recovery plus death) among all infected; 65 + / 15 + is the percentage of the population aged 65 and over from the population aged 15 and over; Date_250 is a dummy variable that takes the value of 1 for the South Korea and adds 1 for each day, of how later the 250th infected person was recorded in the country (for example, for Italy it is 3, for Russia 28); Tests / Population - the number of tests per thousand of the country’s population; Adj. R2 is the corrected coefficient of determination; N is the number of observations. The coefficients are random variables, so after each its standard deviation (standard error) is stated in parentheses. The level of coefficients significance is indicated by asterisks, this is the probability that we consider an insignificant regressor significant: * - 0.1; ** - 0.5; *** - 0.01; **** - 0.001; the absence of asterisks means the probability is more than 10%.

According to the models, the proportion of completed cases has the greatest influence on the detected mortality rate, because, usually, at the epidemic’s beginning, the most severe cases are noticed first.

The next most important factor is the age structure of the population. Should be noted, that here the factor is the proportion of people 65+ not of the entire population, but only of adults, since children, being the infection carriers, along with everyone else, rarely have clinical manifestations of the disease. In Russia too, as a rule, an increased mortality rate is seen not where the general population is older, but where the proportion of older people in the adult population is higher. For example, in the southern republics the population is relatively young, because the share of children is high, but the share of older people in the adult population is also higher than the national average, since life expectancy is higher. In Russia, the value of the index “65+/15+” (17.9) is significantly lower than in Italy (26.3) and Spain (22.7), therefore the lethality is expected to be lower.

The regressor “Date_250” is included in the model with a minus sign, because although there are still no reliable drugs for COVID-19 or a treatment protocol, over time, the global medical community is gaining experience, positive and negative.

In the second model, two factors characterizing the testing level are significant: per one case and per thousand people, both are only 10%, although the importance of testing is undeniable. Model 1 is presented to show that in the absence of one of these regressors, the significance of the other is higher: about 5%. But still this is lower than the revealed significance of some other factors.

There are several reasons for this. First of all, different countries use different tests of different quality, and at the beginning of the pandemic there were no high quality tests at all. Over time, all countries strive to increase testing per case in order to establish the presence or absence of the virus in as many people as possible who may have been in contact with the infected. However, from April 27th to May 7th, this indicator decreased in 27 countries and territories out of 107. In 5 of them this happened because the number of tests done for some reason was not updated on the site (Egypt, Algeria, Guatemala, Mali, Norman Islands). It can be assumed that in the remaining 22, the number of cases is growing faster than the number of completed tests, because either the quality of the initial testing was especially low, or time was lost, and the situation got out of control. The list (in decreasing order of this index decrease in absolute value) is the following: UAE, Ghana, Russia, Kenya, Bahrain, Colombia, Belarus, Nigeria, Bolivia, Brazil, Chile, Honduras, Mayotte, Qatar, Afghanistan, Pakistan, Mexico, Peru, Armenia, Niger, Singapore, Bangladesh. Thus, Russia is among the three worst countries. It is also possible that in some of these countries only narrow selection of the population is repeatedly tested, while testing the rest of the population is not given due attention.

Let us now consider which observations in Model 2 have the largest and smallest residuals with respect to the standard deviation, and try to understand the reasons.

Only Honduras (3.63) goes beyond the three sigma, followed by Belgium (2.93), Sweden (2.22), Hungary (2.16), Bolivia (2.11), Ecuador (1.89), Philippines (1.86), France (1.77), and Indonesia (1.65). The largest negative balances in absolute value are less in absolute value than positive ones: Singapore (−2.31), Qatar (−1.84), Japan (−1.75), Belarus (−1.65), Russia (−1.51), Saudi Arabia (−1.40), Serbia (−1.28), Chile (−1.28), Armenia (−1.19).

Curiously, 8 of these 18 countries were previously listed among the countries with possibly the biggest testing problems. Inadequate testing could lead to both an increase in mortality (Honduras, Bolivia) and an underestimation of the number of real deaths (Russia, Belarus, Chile, Qatar, Armenia, Singapore).

Large positive deviations from the values calculated according to Model 2 could also be caused by both real problems (for example, complete refusal of Sweden from quarantine, mortality outbreaks in nursing homes in France) and by peculiarities of the statistical accounting (for example, in Belgium, to COVID-19 deaths were also added deceased in nursing homes who were suspected of having the disease, even if there was no confirmation^8^). As for the large negative deviations, one should remember that Japan more than once has been praised by the press for its good organization of the measures against the pandemic. Apparently, not in vain, because anyone can go to the local clinic and do a CT scan without any problems.^9^

But do Belarus and Russia have the same level of healthcare? Here it is appropriate to turn to the question of the proportion of medical workers among infected and among the deceased infected. All over the world, health workers are at risk because they come into contact with infected people, while nowhere in the world at the beginning of the pandemic there was a sufficient amount of protective equipment. WHO estimates the proportion of health workers among all infected in the world at about 16%.^10^ Although it varies greatly by country: 20% in Spain, 10% in Italy, 2.8% in Germany, and 1.5% in the USA.^11^ The chances for the medical personnel in Moscow (where the availability of protective equipment is better than in the regions) to become infected are 12 times higher than for other Moscovites.^12^

However, the situation with the proportion of the deceased infected in the world is fundamentally different, because health workers have the opportunity of early disease detection and timely treatment. In many countries, Memory Lists of Deceased Health Workers are being created and this allows us to make a comparison. In Italy, the proportion of medical workers among the deceased infected is 0.6%, in Germany 0.2%, in the United States 0.3%^13^. That is, usually the share of medical workers among the deceased is 5-15 times less than their share among all the infected. But in Russia and Belarus things are different. There were 14 names in the Belarusian Memory List as of May 5th^14^, that is, the share of registered deaths from the epidemic was about 13%. On May 10th, there were 147 names in the Russian List^15^ in 33 regions of the Russian Federation, this is 7.7% of all COVID-19 victims in Russia by that day, of which 47 were in Moscow (4.4% of the deceased), 27 in Dagestan (**150**%) (!!!), 21 in the Moscow region (11%), 14 in St. Petersburg (26%), 5 in the Krasnodar Territory (23%), the remaining 33 deceased health workers accounted for 5.9% of the remaining 563 deceased infected. This indicates both the underestimation of the COVID-19 deceased data in the Russian Federation, and the unsatisfactory quality of the Russian healthcare system.

Thus, the relatively low proportion of COVID-19 deaths in Russia and Belarus is, obviously, primarily the result of a special approach to the cause of death determination. For example, if cancer, atherosclerosis or diabetes complicates the course of the disease resulting from a new coronavirus infection, then in most countries COVID-19 is indicated as the cause of death or one of the causes, and in Russia it is customary to indicate only one main cause, and in these cases it’s usually cancer, acute vascular disease and diabetes^16^. In essence, such definitions of the death causes is contrary to WHO recommendations.

So, to predict the total number of victims of the epidemic in Russia, it is possible to refer to the constructed models from Table 2.

Typically, the number of detected infections declines slowlier than increases. Therefore, if we assume that by May 10th, Russia reached or almost reached the peak of the detected cases per day, then the total number of infected during the pandemic will vary from 750 thousand to a million people. Then the total number of victims of the epidemic can be 14-19 thousand people. However, the standard error of the equations is large due to large errors in the initial data and a small sample. Therefore, the obtained forecast is very approximate.

### Estimation of “additional” mortality rate during the epidemic based on the FSSS demographic data for April

The FSSS published data on fertility and mortality rates in the regions for April only on June 13th, two weeks later than usual, with a note that the data may be incomplete due to the quarantine measures as not all Russians were able to process documents on the births and deaths of their loved ones in April.

In January and February 2020, due to the warm winter, the total number of deaths in the Russian Federation (excluding the Crimea) was 4.9% and 3.7% less than a year earlier. An excess of 0.6% was recorded in March, with a total decline of 2.8% in January-March. Surprisingly, in April, a decline of exactly 2.8% was recorded again (perhaps because not all the dead could be registered). However, in a number of regions, the number of deceased was exceeding even in the April data.

Only four regions saw an increase in the mortality rate in February, March, and April compared to the last year: in Moscow (by 190, 169 and 1934, respectively), Moscow region (258, 217 and 1018), Penza region (25, 87 and 52) and Chuvashia (71, 47, 34). In the other 6 regions, an excess of mortality rate was recorded only in February and March (probably, in April, not all deaths were registered), these are Vladimir region (by 65 and 15), Pskov region (69 and 23), Tambov region (24 and 95) and Ulyanovsk region (143 and 125), Karelia (9 and 148) and KhMAD (3 and 110), a similar picture was observed in Sevastopol (9 and 7). In YNAD, the excess in the mortality rate was observed in February and April (by 9 and 23). In the other 5 regions, the increase in the mortality rate happened in March and April, these are Leningrad region (by 253 and 116), Lipetsk region (139 and 40) and Tomsk region (72 and 172), Altay (12 and 20) and Khabarovsk Territory (28 and 92). In the 24 regions of the Russian Federation, the excess of the mortality rate has been recorded only in March, these are Vologda (by 26), Kaluga (57), Kemerovo (275), Kurgan (25), Murmansk (3), Nizhny Novgorod (275), Novgorod (42), Novosibirsk (73), Oryol (92), Samara (142), Saratov (2), Sverdlovsk (443), Tula (109), Tyumen without autonomous districts (17) and Yaroslavl (91) regions, the Republic of Buryatia (55), Ingushetia (2), Mari El (51), Tatarstan (211) and Udmurtia (72), Zabaikalsky (133), Krasnoyarsk (110), Perm (96) and Primorsky (49) regions. In 8 other regions of the Russian Federation, the excess in the mortality rate was registered only in April, these are St. Petersburg (by 114), Belgorod (23), Bryansk (3), Kaliningrad (22), Kirov (4), Kostroma (108) and Tverskaya (8) regions and the Kabardino-Balkarian Republic (4).

Of course, not all of these additional deaths are associated with the coronavirus, especially in February. Both in February and March, the pandemic, apparently, had only a small, albeit significant, effect on the mortality rate in the Russian Federation: the correlation coefficient of changes in the mortality rate in February and March (in total) in 2020 compared to the same period of the previous year, with the people infected with the new coronavirus on March 31st, resulted in 0.300, and with the number of officially registered Covid-19 deaths - 0.277. The addition of April values changes the picture dramatically: the correlation of changes in the mortality rate from all causes in February-April compared to the same period of 2019 with the number of infected as of April 30th is already 0.743, and with the number of Covid-19 deaths in the regions of Russia - 0.715. The correlation coefficient of only April mortality changes with the number of infected is 0.789, and with the number of Covid-19 deaths - 0.765. Should be noted that in all the cases, the correlation with the number of infected is higher than with the number of Covid-19 deaths. This is further evidence that the official coronavirus mortality statistics are not very close to reality.

The second side of the issue is the April mortality undercount. This could have happened even in the regions listed above. To figure out where it was most significant, there are three ways: 1) to create the regression equations and to see where are the smallest (i.e., the largest negative) residuals of these equations; 2) to compare the change in birth and mortality rates in April YoY, because registration problems with the births and deaths were most likely in the same subjects of the Russian Federation; 3) to analyze the statistics of the diseased infected health workers.

1. Let us find the residual of the two regression equations

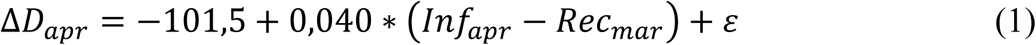

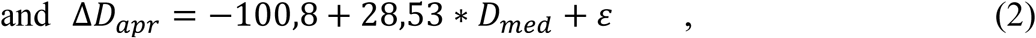

where Δ*D*_*apr*_is the change in the mortality rate in April 2020 compared to April 2019, *Inf*_*apr*_is the number of people infected with the new coronavirus by April 30th in the regions, *Rec*_*mar*_is the the number of Covid-19 recoveries by March 31st, *D*_*med*_is the number of infected health workers who died before May 10th, *ε* is the equations residual; The variables coefficients were found empirically from 83 observations, the determination coefficient is R^2^=0.624 for the first equation and 0.368 for the second. There are two equations, because it is impossible to include both explained variables in one equation, since there is a very high correlation between them, 0.821. The number of infected deceased health workers adds to the idea of the epidemiological situation in the regions, since their share in the total number of infected people is higher in places where the severe cases proportion is higher among the infected, and the medical care organization is the least satisfactory. Undoubtedly, it would be better to include in the equation the number of infected health workers who died during April, but the Russian Memorial List does not state the death dates, and the authors do not have earlier data than of May 10th. The first equation states that, on average, 4 out of every 100 infected people died in April, while the second suggests that 28 other people died for every infected deceased health worker. According to the first equation, mortality is most underestimated in **Krasnodar Territory** (residual is −1040), **Dagestan** (−563), **Irkutsk** (−387) and Rostov (−338) regions, **Stavropol Territory** (−261), Sverdlovsk (−228), **Ulyanovsk** (−190),Tula (−157) regions, **Altay Territory** (−135), **Saratov region** (−134), **Zabaykalsky Territory** (−132), **Pskov region** (−131) and **Chechen** R. (−118). According to the second equation - in **Dagestan** (−1278), **Krasnodar Territory** (−1143), **Irkutsk** (−383) and Rostov (−330) regions, **Stavropol Territory** (−296), **Ulyanovsk** (−203) and Sverdlovsk (−200) regions, **St. Petersburg** (−185), Tula Region (−148), **Zabaykalsky Territory** (−128), **Chechen Republic**. (−128) and **Pskov region** (−126). Unfortunately, the equations do not show how much the mortality rate in April is underestimated, since the sum of all values calculated from the equation is always equal to the sum of the initial values of the explained variable. The second way makes the situation a little clearer.
2. If we follow the second path, it turns out that, perhaps, in most regions in April, not all births and/or deaths were registered (in Table 4 only regions where the value in at least one of columns 6 and 7 does not exceed −5 are marked). This is especially true for such regions as the republics of **Dagestan**, Ingushetia, Kalmykia, **Chechen**; Pskov, **Ulyanovsk, Irkutsk** and Sakhalin regions, **Krasnodar** and Zabaykalsky territories. Of the regions for which the largest negative residuals in the equations were observed, Table 4 does not contain just Rostov and Sverdlovsk regions.
3. A little more clarity can be added by analyzing the mortality statistics of the infected health workers. It was already mentioned that the largest share of the deceased health workers among the infected was on May 10th in **Dagestan, St. Petersburg** and the **Krasnodar Territory**, and the largest absolute values were in Moscow and the Moscow Region. If we assume that the majority of additional deaths in February-April in Moscow and the Moscow region are associated with the pandemic, then in these two subjects of the Russian Federation the proportion of doctors among the deceased infected is only 2%. In other regions, this share is probably higher, since the state of the healthcare system is worse. Thus, the coefficient on the variable “health worker deaths” in equation 2 appears to be close to reality. **Therefore, the total number of additional deaths in April should be close to 4**.**5 thousand, and the total number of deaths from all causes was probably about 3% higher in April 2020 than in April 2019**.

### Estimation of the “additional” mortality during the pandemic based on demographic data from the RSSS for April-May

The RSSS also released data on the birth and mortality rates in the regions for May two weeks later than usual, on July 14th, with the same note that the data may be incomplete due to the quarantine measures. At the same time, updated data for April were released. However, apparently, some part of April deaths in more regions got into the May statistics. This is proved by the following: in preliminary data, the number of births in April 2020 on the territory of the Russian Federation (excluding the Crimea) amounted to 91.9% in 2019, and the number of deaths was 97.2%, and according to the updated data, 94.7% and 98.2% respectively. That is, the number of births was adjusted by 2.9 percentage points, and the number of deaths by only 1.0 percentage points. At the same time, in the preliminary data for May, the number of births was only 90.1% of 2019, and the number of deaths is 112.4%.

**Table 4.** Change in the number of birth and mortality rates in January-March and April YoY

| Region | Number of births,<br>2020 in % in 2019 |  | Number of deaths,<br>2020 in % in 2019 |  | Changes |  |
| --- | --- | --- | --- | --- | --- | --- |
|  | January-March | April | January-March | April | (3)-(2) | (5)-(4) |
| 1 | 2 | 3 | 4 | 5 | 6 | 7 |
| Dagestan R. | 89,9 | 80,2 | 99,9 | 48,9 | -9,7 | **-51,0** |
| Chechen R. | 108,4 | 98,0 | 99,7 | 71,7 | -10,4 | **-28,0** |
| Nenets A.D | 100,0 | 115,3 | 113,1 | 87,9 | 15,3 | **-25,2** |
| Pskov region | 87,5 | 100,4 | 104,2 | 79,8 | 12,9 | **-24,4** |
| Ulyanovsk region | 96,2 | 95,9 | 105,5 | 83,1 | -0,3 | **-22,4** |
| Ingushetia R. | 97,0 | 53,5 | 93,9 | 74,6 | **-43,5** | **-19,3** |
| Krasnodar t. | 93,7 | 88,7 | 101,1 | 82,7 | -5,0 | **-18,4** |
| Zabaykalsky t. | 96,9 | 91,1 | 100,1 | 82,0 | -5,8 | **-18,1** |
| Kalmykia R. | 100,6 | 83,8 | 98,1 | 81,0 | **-16,8** | **-17,1** |
| Irkutsk region | 94,7 | 98,9 | 101,2 | 84,7 | 4,2 | **-16,5** |
| Karelia R. | 93,0 | 83,8 | 103,3 | 89,7 | -9,2 | -13,6 |
| Mari El R. | 98,3 | 95,5 | 100,2 | 87,8 | -2,8 | -12,4 |
| Tula region | 94,4 | 101,3 | 100,8 | 89,1 | 6,9 | -11,7 |
| Novgorod region | 91,2 | 87,4 | 102,3 | 90,6 | -3,8 | -11,7 |
| Sakhalin region | 100,1 | 83,3 | 96,8 | 85,8 | **-16,8** | -11,0 |
| Adygeya R. | 109,4 | 110,7 | 93,1 | 82,6 | 1,3 | -10,5 |
| Orlovsky region | 95,7 | 90,9 | 97,9 | 87,8 | -4,8 | -10,1 |
| Mordovia R. | 89,5 | 102,7 | 97,8 | 87,9 | 13,2 | -9,9 |
| Crimea R. | 96,8 | 98,6 | 100,1 | 91,2 | 1,8 | -8,9 |
| Magadan region | 99,1 | 106,3 | 94,1 | 85,4 | 7,2 | -8,7 |
| Kaluga region | 94,3 | 94,7 | 98,1 | 90,6 | 0,4 | -7,5 |
| Khakassia R. | 93,6 | 86,7 | 98,1 | 90,7 | -6,9 | -7,4 |
| Saratov region | 90,6 | 92,0 | 99,2 | 92,1 | 1,4 | -7,1 |
| Karachayevo-Cherkessian R. | 97,4 | 89,2 | 96,6 | 90,1 | -8,2 | -6,5 |
| North Ossetia R. | 92,7 | 85,5 | 93,6 | 87,1 | -7,2 | -6,5 |
| Kamchatka t. | 97,7 | 89,5 | 98,8 | 92,4 | -8,2 | -6,4 |
| KhMAD | 98,3 | 86,2 | 103,4 | 97,1 | -12,1 | -6,3 |
| Komi R. | 96,7 | 85,0 | 95,5 | 89,8 | -11,7 | -5,7 |
| Astrakhan region | 98,0 | 89,1 | 101,5 | 96,5 | -8,9 | -5,0 |
| Stavropol t. | 95,0 | 95,0 | 92,5 | 87,5 | 0,0 | -5,0 |
| Lipetsk region | 90,9 | 84,4 | 99,2 | 102,8 | -6,5 | 3,6 |
| Yaroslavl region | 92,3 | 86,5 | 97,7 | 99,9 | -5,8 | 2,2 |
| Kemerovo region | 94,1 | 88,5 | 93,6 | 98,5 | -5,6 | 4,9 |
| St. Petersburg | 95,8 | 90,4 | 98,5 | 102,2 | -5,4 | 3,7 |
| Volgograd region | 91,9 | 86,7 | 96,1 | 98,6 | -5,2 | 2,5 |
| Leningrad region | 95,4 | 90,2 | 102,8 | 105,9 | -5,2 | 3,1 |
| Murmansk region | 102,0 | 96,9 | 101,1 | 99,6 | -5,1 | -1,5 |
| Altay R. | 92,8 | 87,8 | 91,6 | 92,2 | -5,0 | 0,6 |

The birth rate was adjusted in 7 constituent entities of the Russian Federation (the difference in percentage points is stated in brackets): in the Republic of Ingushetia (by 47.1), Dagestan (34.4), Chechen Republic (26.9), Yakutia (9.7), Moscow (8.5), St. Petersburg (5.0) and Karachay-Cherkess Republic. (4.0).

The April mortality rate was adjusted in 8 regions: the republics of Dagestan (55.4), Ingushetia (38.1), Chechen (20.2), North. Ossetia (12.1), Kabardino-Balkaria (8.4), Adygea (7.5), Karachay-Cherkessia (6.3) and Krasnodar Territory (5.7). Perhaps, in some cases, the birth rate really needed a greater adjustment than the mortality rate, for example, in Moscow.

As for the May’s mortality rate data, they are bigger than in 2019, in 59 constituent entities of the Russian Federation (excluding the Crimea) out of 85. The largest increase in the mortality rate YoY was recorded in the republics of Ingushetia (by 69.4%) and Dagestan (by 59.4%), followed by Moscow (by 57.2%), St. Petersburg (by 46.7%), Chechen Republic (by 44.9%), and Moscow region (by 44.1%), North Ossetia (by 42.5%), Mordovia (by 41.3%), YNAD (by 37.6%), Leningrad region (by 29.5%), Penza region (by 26.4%), Kabardino-Balkarian R. (by 23.7%), Chukotka (by 23.7%), Saratov (by 19.8%), Smolensk (by 19.7%), Kaluga (by 19.5%) and Tula (by 19.2%) regions.

Earlier we assumed (on the basis of Table 4) that in most of these regions (the republics of Adygea, Dagestan, Ingushetia, Karachay-Cherkessia, Mordovia, North Ossetia, Chechnya, Krasnodar Territory, St. Petersburg, Leningrad, Saratov, Kaluga and Tula regions) the mortality rate was underreported, now we see arguments in favor of the fact that these assumptions were correct.

The mortality rate data in April were not corrected in all the regions, where this should have been done, and in a number of regions, part of the April values was apparently attributed to May. At the same time, some of the May deaths were also not registered, as stated in the RSSS data notes. To assess which regions most likely had the most significant underestimation, we will follow the same three ways described in the previous section. To begin with, let’s build table 5, similar to Table 4, with one difference: all regions with a negative value in column 7 are included here, since in May it was impossible to say that the pandemic peak had passed about any region, and in column 6, as before, we will consider only values less than −5 as doubtful. Although in some cases (for example, in Moscow), a significant decrease in the birth rate may be due not to the data underestimation, but rather to the outflow of migrants, both external and internal.

**Table 5.**
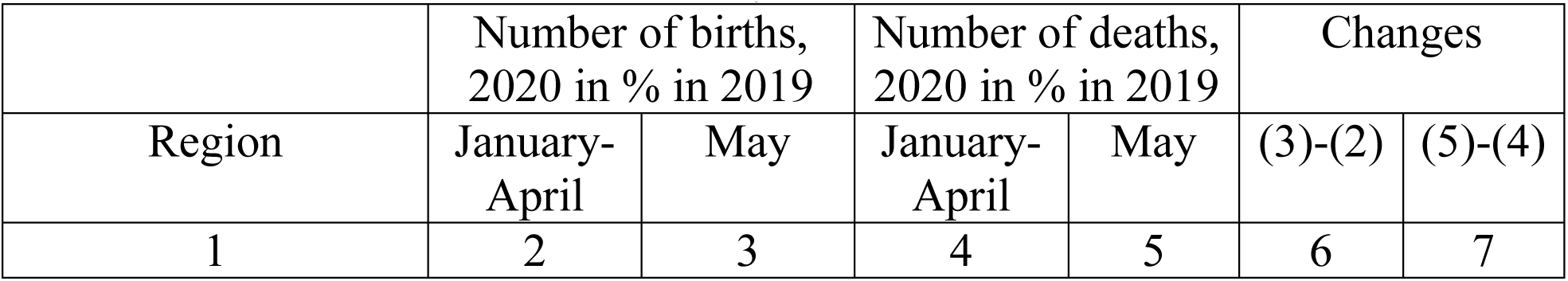
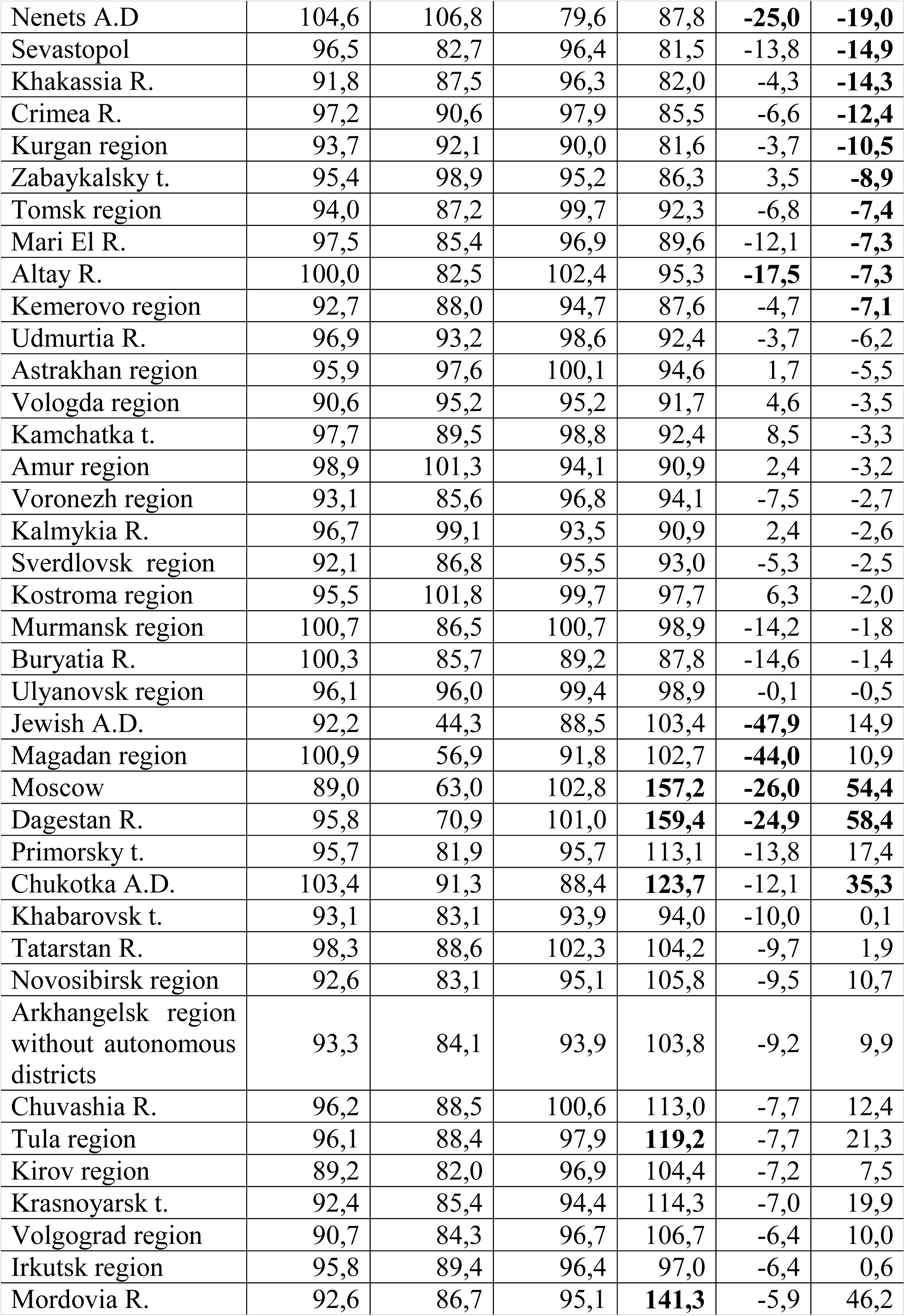
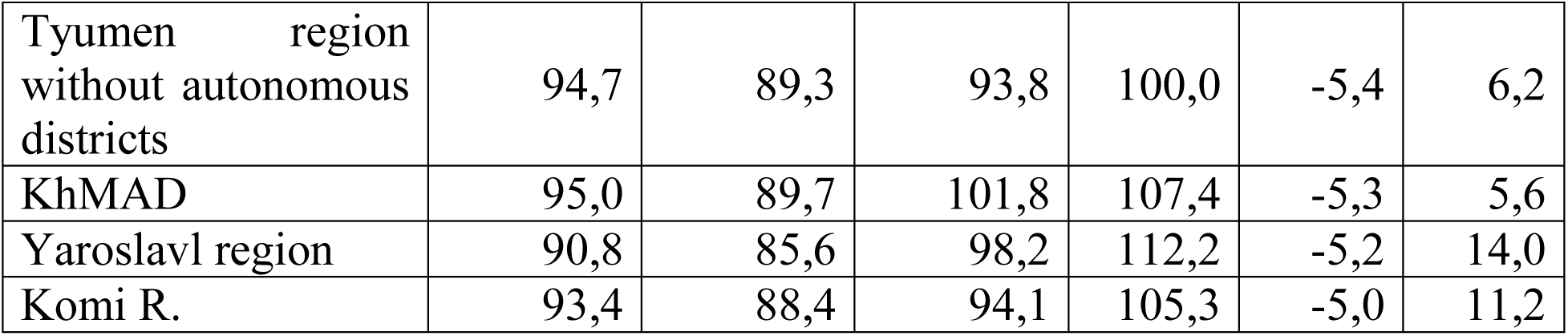
Change in the number of birth and mortality rates in January-April and May YoY

If we compare the values in Tables 4 and 5, it can be assumed that in some other regions (for example, in the Yaroslavl region and Krasnoyarsk Territory) a part of the April deaths were recorded only in May.

Thus, in economic modeling, it is better to take the change in the sum of the number of deaths in April and May as an explained variable:

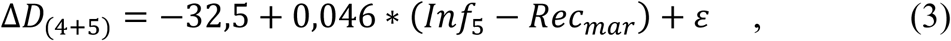

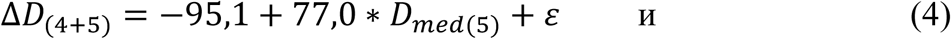

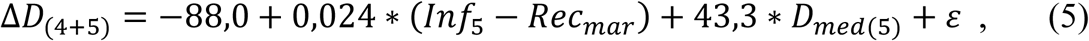

where Δ*D*_(4+5)_is the change in the mortality rate in April and May 2020 compared to April-May 2019,

*Inf*_5_ is the number of people infected with the new coronavirus by May 31st in the regions,

*Rec*_*mar*_ is the number Covid-19 recoveries from by March 31st,

*D*_*med*(5)_ is the number of infected health workers who died before June 1st,

*ε* is the equations residual.

The variables coefficients were found empirically from 83 observations, the determination coefficient is R^2^=0,831 for the equation (3), 0,842 for the equation (4), and 0,912 for the equation (5), thus, the relationship between excess mortality and the selected regressors is now much greater than it was according to the incomplete April data.

Now, it became possible to construct model (5) with two regressors, because the correlation of each of the regressors with the explained variable is greater than with each other. Perhaps this happened because now the information on the number of people infected in the regions is less reliable than on the number of “additional” deaths. This is also indicated by the comparison of the remainders of the equations (3) and (4). Apparently, the record holder for underestimating the number of infected among the regions of the Russian Federation is Dagestan, since it has the smallest residual in equation (4), −2788, and one of the largest residues in equation (3).

The data on the deceased infected health workers are the most reliable, since each, thanks to the Memory List, is known by name. But, fortunately, not all the regions have the doctors who died during the fight against the pandemic, so both regressors complement each other.

The smallest (largest negative) residuals in equation (5) were in Dagestan (−1301), Sverdlovsk (−731) and Irkutsk (−519) regions, Altai Territory (−415), Kemerovo (−395), Chelyabinsk (−378) and Ulyanovsk (−350) regions, Zabaykalsky Territory (−322), Moscow (−307) and Kursk region (−300). Table 5 does not include the Altai territory, Chelyabinsk and Kursk regions. However, 8 infected doctors died in Kursk region, so the data on mortality there is most likely underestimated. A negative residuals in Moscow does not necessarily mean that data on mortality is underestimated. This may be due to a better health organization than in other regions.

## Conclusion

The data for April and May revealed 15,381 additional deaths compared to 2019. Of these, according to official data from the website CORONAVIRUS (COVID-19)^17^, which are used in international comparisons, as of May 31^st^, only 4,699 people died due to COVID-19. According to data from FSSS, which were more complete, 6994 people died in April and May with a proven diagnosis COVID-19 as the main cause of death. 2198 deaths could have COVID-19 as the main cause of death, but the virus has not been identified, and 6085 deaths were not caused by COVID-19 itself but the virus was referred to “other important conditions”. Thus, 15,277 additional deaths in April and May were associated with the coronavirus.

At the end of May, it was known about 405843 infected people and 311 dead doctors, as well as about 120 recovered on March 31^st^. It is known that by May 31st 171,883 people have recovered, while by July 31st, the total number of infected people in the Russian Federation was 839,981, and the total number of the deceased infected health workers reached 620. Applying the formula (5), we can conclude that the number of direct and indirect pandemic victims in the Russian Federation at the end of July amounted to approximately 43 thousand people. However, most likely, this number is overestimated, since medicine over time better copes with new challenges, so the coefficient of the variable reflecting the number of patients should gradually decrease. If we focus only on the number of deceased doctors, then the number of additional deaths should be approximately 31 thousand.

This number may be less if part of the deaths in April-May was compensatory after the favorable January and February. However, it may be more than 31 thousand, if the proportion of deceased doctors among all the dead infected has decreased over time. In addition, there was also some underreporting of deaths in May, as in the preliminary data for April.

Thus, our initial estimation of 14-19 thousand victims of the epidemic in Russia based on the global data by May 7th, which at that time seemed quite high against the background of the official COVID-19 death toll, in reality turned out to be too optimistic.

## Data Availability

Data openly available

https://www.worldometers.info/coronavirus/

## Footnotes

2 https://promdevelop.ru/science/statistika-smertnosti-v-rossii-v-usloviya-koronavirusa/

3 www.gks.ru

4 https://databank.worldbank.org/home.aspx

5 https://coronavirus-monitor.ru/coronavirus-v-rossii/

6 https://www.worldometers.info/coronavirus/

7 https://www.who.int/classifications/icd/Guidelines_Cause_of_Death_COVID-19.pdf

8 https://rg.ru/2020/04/22/obiasnen-samyj-vysokij-v-mire-uroven-smertnosti-ot-covid-19-v-belgii.html

9 https://www.bbc.com/russian/vert-tra-52404096?fbclid=IwAR00rKby1S7I5l4NgvNhwlYqm0YW9fC34pmhsnu3EExedEcqdarFNZwPbrs

10 https://www.bbc.com/russian/features-52493773; https://www.bbc.com/russian/features-52112233

11 https://newsmaker.md/rus/novosti/v-moldove-dolia-zarajhennyh-vrechei-odna-iz-samyh-vysokih-v-mire-covid-19-v-tsifrah/

12 https://www.vedomosti.ru/society/articles/2020/05/01/829484-ne-hvataet-sredstv-zaschiti?utm_campaign=editorchoice09052020&utm_medium=email&utm_source=newsletter

13 https://gomel.today/rus/article/society-1558/

14 https://www.svaboda.org/a/30582868.html

15 https://sites.google.com/view/covid-memory/home?fbclid=IwAR0ouB2JGH2MFF7nKF6zkKtS4jxcgWOpWieGTXfX7mPXiFSHQqJVD1-ZsMA

16 New coronavirus infection (COVID-19). Rules of work of pathology departments. Temporary recommendations. Version 2. Moscow, April 27, 2020. Pages 23-28. http://patolog.ru/sites/default/files/metodichka_dz_.pdf

17 https://coronavirus-monitor.ru/

## Notes

### Competing Interest Statement

The authors have declared no competing interest.

### Funding Statement

No funding was received at any stage of conducting this research.

